# Sex differences, cross-ancestry generalizability, and noise-smoking interactions in the polygenic architecture of hearing loss in adults

**DOI:** 10.1101/2022.01.26.22269898

**Authors:** Flavio De Angelis, Oana A. Zeleznik, Frank R. Wendt, Gita A. Pathak, Daniel S. Tylee, Antonella De Lillo, Dora Koller, Brenda Cabrera-Mendoza, Royce E Clifford, Adam X. Maihofer, Caroline M. Nievergelt, Gary C. Curhan, Sharon G. Curhan, Renato Polimanti

## Abstract

We conducted a comprehensive genome-wide investigation of hearing loss (HL) in 748,668 adult participants of the UK Biobank, the Nurses’ Health Studies (I and II), the Health Professionals Follow-up Study, and the Million Veteran Program. We identified 54 risk loci and characterized HL polygenic architecture, exploring sex differences, polygenic risk across ancestries, tissue-specific transcriptomic regulation, cause-effect relationships with genetically-correlated traits, and gene interactions with HL environmental risk factors. Our transcriptomic regulation analysis highlighted the potential role of the central nervous system in HL pathogenesis. This was supported by the multivariate interaction analysis that showed how genes involved in brain development interact with sex, noise pollution, and tobacco smoking in relation to their HL associations. Additionally, the genetically-informed causal inference analysis showed that HL is linked to many physical and mental health outcomes. These results provide many novel insights into the complex biology and epidemiology of HL in adults.

## INTRODUCTION

Acquired hearing loss (HL) is the third most common chronic health condition^1^ and the fourth leading cause of disability globally^2^. The World Health Organization (WHO) report of hearing projects that nearly 2.5 billion individuals will have some degree of HL by 2050^3^. HL is associated with several comorbid conditions^4^. For instance, HL-induced impaired communication ability particularly among older people can lead to social isolation with major health, psychosocial, and economic consequences, reducing the quality of life^4, 5^. HL affects individuals of all ages, but its prevalence increases with age, reflecting the cumulative effect of environmental factors and genetic predisposition^6^.

Heritability estimates for acquired HL range from 30% up to 70%^7^, highlighting that genetic variation is a key determinant for individual HL risk. More than 100 genes present mutations that result in congenital HL not associated with disorders in other organs or dysmorphic features (non-syndromic HL)^8, 9^. Mutations causing congenital HL affect genes involved in cochlear function, specifically affecting the sensory and mechanosensory cells^10–12^. Beyond these Mendelian forms, acquired HL cases appear to be due to the additive effect of many common genetic variants with small individual effects. Large-scale genome-wide association studies (GWAS) conducted in population-based cohorts have identified more than 50 common risk variants and characterized the regulatory role of these loci in multiple cell and tissue types^13–16^. Although these studies have generated important insights into the genetic predisposition to HL, there are several aspects of the HL pathogenesis that are currently under-investigated by genetic studies. For instance, HL among older adults is more common, more severe, and with earlier onset in men than in women, even after adjusting for confounding factors such as higher occupational noise exposure in men^17, 18^. However, the molecular pathways that underlie HL sex differences are unclear. Similarly, we have a limited understanding which are the biological processes interacting with HL environmental risk factors. Several studies showed that noise pollution and tobacco smoking are HL risk factors^19–22^, but to date no large-scale studies investigated how genetic variation interacts with noise pollution and tobacco smoking in determining HL risk.

In the present study, we conducted an extensive genome-wide investigation across the UK Biobank (UKB; 251,233 women and 214,549 men)^14^, the Nurses’ Health Studies (NHS I, 14,978 women; and NHS II, 12,533 women)^23, 24^ and the Health Professionals Follow-up Study (HPFS, 8,532 men)^25^. The risk loci identified were replicated in a sample of 226,043 participants from the Million Veteran Program (MVP)^26^. Our findings provide a more comprehensive understanding of the genetic basis of HL sex differences, uncovering novel sex-specific risk loci, molecular processes, putative cause-effect relationships, and the interaction of genetic variation with sex, noise pollution, and smoking behaviors.

## METHODS

### Cohorts and hearing-loss assessment

We leveraged genome-wide information from 748,668 adult participants recruited by the UKB, NHS I, NHS II, HPFS, and MVP cohorts to investigate the polygenic architecture of HL. UKB is a large population-based research resource, containing in-depth genetic and health information from over 500,000 UK participants 40-69 years at enrollment^14^. UKB HL-related phenotypes were defined by self-reported items derived from a touchscreen questionnaire and audiometric measurements assessed through the Speech Recognition Threshold (SRT) test. The self-reported items included the following binary traits: *“Do you have any difficulty with your hearing?”* (UKB Field ID: 2247); *“Do you find it difficult to follow a conversation if there is background noise (such as TV, radio, children playing)?”* (UKB Field ID: 2257); and *“Do you use a hearing aid most of the time?”* (UKB Field ID: 3393). UKB participants who indicated they were completely deaf (N=144) were excluded from the analysis to reduce the likelihood of including congenital forms of HL^16^. For ∼12% of the UKB participants (59,807 for UKB Field ID 2247; 60,448; and 40,656 for UKB Field ID 3393), these items were assessed multiple times. Since age is a strong risk factor for acquired HL, we considered the most recent assessment when multiple assessments were available to improve the ability to detect the disease onset. In addition to considering these binary traits individually, we combined them in a four-category ordinal phenotype (Supplemental Table 1). For the SRT-derived audiometric measurements, we considered the UKB item “*the signal-to-noise ratio at which half of the presented speech can be understood correctly”* for both left and right ears (UKB Field ID: 20019 and 20021, respectively). Because of the much larger sample size, we included unrelated UKB participants of European descent in the primary discovery sample. The other ancestry groups available in UKB were used for single-variant and polygenic risk score (PRS) replication (see section Cross-ancestry Replication and Polygenic Risk Scoring).

Additional genome-wide information was derived from NHS I, NHS II, and HPFS cohorts. The NHS I began in 1976 when 121,700 female registered nurses, aged 30–55 years, were enrolled by completing a baseline questionnaire about their health and lifestyle. In 1989, the NHS II was established and enrolled 116,429 younger female registered nurses, aged 25 to 42 years^24^. The HPFS began in 1986 and enrolled 51,529 male health professionals, aged 40 to 75 years. In each of the cohorts, detailed information on demographics, health, diet, and lifestyle factors was collected and updated every 2 years (every 4 years for diet). The follow-up rates in all 3 cohorts exceed 90% of eligible person-time^27^. Self-reported hearing status was determined based on participants’ responses to biennial questionnaires. *“Hearing difficulty”* was defined as a participant report of a hearing problem that was mild, moderate, severe (non-hearing aid user), or severe (hearing aid user). Leveraging samples with genome-wide information, we investigated 14,978 unrelated female NHS I participants, 12,533 unrelated female NHS II participants, and 8,532 unrelated male HPFS participants, all of European descent. To maximize the sample size available, we meta-analyzed NHS I and II cohorts and referred to them hereafter as a single NHS sample.

The MVP is a biobank funded by the US Department of Veterans Affairs that to date enrolled more than 800,000 participants among active users of the Veterans Health Administration healthcare system^26^. In our analysis, we used genome-wide information regarding 85,743 cases and 140,300 controls based on self-reported hearing-loss information. Information regarding HL assessment in MVP has been previously described ^28^.

### Genome-wide data quality control and GWAS meta-analysis

We used UKB imputed data ^29^, filtering variants with imputation score < 0.8, minor allele frequency (MAF) < 0.01, Hardy-Weinberg Equilibrium p-values <10^-6^, and missingness > 0.1. The genetic data were employed to identify related individuals by estimating kinship coefficients for all pairs of samples. Accordingly, we removed individuals based on relatedness (one individual from pairs with kinship coefficient > 0.042 was removed, with the preference to retain cases), forming a maximally unrelated subset^29^. In the NHS and HPFS cohorts, SNPs with imputation score <0.8 or minor allele frequency < 0.05 were excluded. Quality checks included the exclusion of those participants with a poor genotype call rate (<95%) and a check for relatedness. The SNPs with a poor call rate (<95%), out of Hardy-Weinberg equilibrium (P<10^-5^), with high duplicate discordance rates, or monomorphic were excluded. Genotyped data from each of the NHS and HPFS studies were imputed using the 1000 Genomes Project Phase 3 reference panel ^30, 31^. Details regarding genotyping and imputation of the MVP samples were described elsewhere^32^. Relatedness was estimated using KING^33^. For each pair of subjects with an estimated kinship coefficient > 0.088 (2nd degree or closer), one individual was removed, with the preference to retain cases. If individuals had the same diagnostic status, one individual was removed at random. HARE (harmonized ancestry and race/ethnicity) estimates^34^ were used to select European ancestry individuals. GWAS were carried out in the UKB, NHS, and HPFS by logistic regression, using PLINK 2.0 ^35^ and including age and the first 10 within-ancestry PCs as covariates. For the sex-combined analyses, sex was also included as a covariate. The GWAS generated by the individual cohorts were meta-analyzed using the inverse variance-based method implemented in METAL^36^. In MVP, GWAS was performed using logistic regression of the hearing phenotype on imputed SNP dosages including covariates for 10 PCs, age, and sex, using PLINK 1.9 software^35^.

### SNP-based heritability and genetic correlation

SNP-based heritability (SNP-h^2^) and genetic correlation (rg) for all hearing traits were estimated using the Linkage Disequilibrium Score Regression (LDSC) method^37^. As recommended by the LDSC developers (details at https://github.com/bulik/ldsc), the analysis was conducted considering the HapMap 3 reference panel and pre-computed LD scores based on the 1000 Genomes Project reference data for individuals of European ancestry. SNP-h^2^ and genetic correlations were evaluated among HL trait assessed in the UKB, NHS, HPFS, and their meta-analyses. Since functional categories of the genome contribute disproportionately to the heritability of complex diseases^38^, SNP-h^2^ partitioning was conducted with LDSC using 95 baseline genomic annotations^39^ such as allele frequency distributions, conserved genomic regions, regulatory elements, and annotations for genic, loss-of-function (LoF) intolerant, positively and negatively selected regions^40^. We also conducted a UKB phenome-wide genetic correlation analysis of HL, testing 7,153 phenotypes for the sex-combined investigation, 3,287 phenotypes for the female-specific investigation, and 3,144 phenotypes for the male-specific investigation. The genome-wide association statistics for the sex-combined analysis were derived from the Pan-UKB data release (available at https://pan.ukbb.broadinstitute.org/downloads). The sex-specific genome-wide association statistics were derived from a previous UKB analysis (available at http://www.nealelab.is/uk-biobank).

### Latent causal variable analysis

To evaluate whether the genetic correlations of HL are due to cause-effect relationships, we used the latent causal variable (LCV) method to conduct a genetically-informed causal inference analysis^41^. As recommended, only SNPs with MAF > 5% were considered, and the major histocompatibility region was removed. Considering traits that reached at least a nominally significant genetic correlation (p<0.05) with HL, we tested 879 traits in the sex-combined analysis, 323 traits in the female-specific analysis, and 332 traits in the male-specific analysis. For each comparison, the genetic causality proportion (gcp) can range from zero (no partial genetic causality) to one (full genetic causality).

### Variant prioritization, fine-mapping, and multi-tissue transcriptome-wide association study

To identify the causal loci underlying the statistical association observed, we functionally annotated GWAS findings and prioritized the most likely causal SNPs and genes using pre-calculated LD structure based on 1000 Genomes Project EUR reference populations^42^. The risk loci identified in the sex-combined and sex-specific meta-analyses were classified considering the following parameters derived from Functional Mapping and Annotation of Genome-Wide Association Studies (FUMA)^42^: leadP = 5×10^-8^; gwasP = 0.05; R^2^ = 0.1; refpanel = 1000 Genomes Project Phase3 EUR reference populations; MAF = 0.01; refSNPs = 1; mergeDist = 250. Positional mapping was performed considering MapWindowSize = 10 and the minimum Combined Annotation Dependent Depletion (CADD)^43^ score for SNP filtering was set to 0. Gene-based analysis and gene-set analysis were performed with Multi-marker Analysis of GenoMic Annotation (MAGMA)^44^ integrated into FUMA. Tissue enrichment analysis was performed using Genotype-Tissue Expression (GTEx) project v8 data^45^.

We fine-mapped the association statistics for a 3MB region around the lead SNP of the genomic risk locus identified by FUMA (r^2^ = 0.1; window = 250bp). Each region for the respective association was fine-mapped to determine 95% credible set using susieR^46^ with at most 10 causal variants (default). The credible set reports variants most likely to be causal based on the marginal posterior inclusion probability (PIP) ranging from 0 to 1, with values closer to 1 to be most causal.

To further explore transcriptomic regulation in the context of HL, we performed a multi-tissue TWAS (transcriptome-wide association study), using the S-MultiXcan approach to combine information across 49 GTEx tissues adjusting for tissue–tissue correlation^47^.

### Cross-ancestry replication and polygenic risk scoring

We used data from the Pan-UKB data release (available at https://pan.ukbb.broadinstitute.org/downloads) to conduct cross-ancestry replication and PRS analyses. Specifically, we derived PRS of HL associations from EUR ancestry (base dataset) and tested in UKB participants of African (AFR N=6,636), admixed American (AMR N=980), Centra/South Asian (CSA N=8,876), East Asian (EAS N=2,709), and Middle Eastern (MID N=1,599) ancestries (target datasets). PRS analysis based on HL meta-analyzed and sex-stratified GWAS was computed using PRSice v. 2.3.1.c^48^, using the clumping-thresholding method to maximize the predictive ability of the derived polygenic scores^49^. Using 1000 Genomes Project EUR populations as reference panel, SNPs were clumped based on 250kb windows, based on clump-r2 threshold=0.1 and clump-p threshold=1, respectively. The step size of the threshold was set to 5×10^-5^, and the range of p-value thresholds was from 5×10^-8^ to p=1 using an additive model for regression at each threshold.

### Multivariate gene-by-environment genome-wide interaction study

A multivariate gene-by-environment genome-wide interaction study (GEWIS) was performed using StructLMM^50^, a linear mixed-model approach to efficiently detect interactions between loci and multiple potentially correlated environments. Since StructLMM assumes a quantitative trait, the GEWIS was conducted considering the four-category ordinal trait described above (Supplemental Table 1). The multivariate GEWIS was performed considering a total of 14 environments, including sex, smoking behaviors and exposures, and different types of noise pollution exposure (Supplemental Table 2). Marginal log-likelihoods (log (Bayes factor -BF)) between the full model and the reduced model with environments removed were used to identify the most relevant environments for the detected locus interaction effects. A pathway enrichment analysis was conducted considering loci with nominally significant multivariate gene-environment interaction. These variants were clumped using R ld_clump function (ieugwasr R package) using clump_kb = 10,000, clump_r2 =0.001, and clump_p =0.05. The resulting variants showing BF>1 were mapped to genes through Ensembl platform^51^. A Gene Ontology (GO) enrichment analysis was performed using the Database for Annotation, Visualization and Integrated Discovery (DAVID)^52^ and GO terms surviving false discovery rate (FDR) multiple testing correction (q<0.05) were retained as significant.

## RESULTS

### SNP-based heritability and genetic correlation among hearing-loss traits

We investigated the SNP-h^2^ across multiple HL traits. In the UKB, these included phenotypes derived from the touchscreen questionnaire and an automated hearing assessment (i.e., SRT test). For the questionnaire-derived HL traits, the SNP-h^2^ ranged from 0.017±0.002 for “*Hearing aid use*” to 0.050±0.002 for “*Hearing difficulties with background noise*”. The genome-wide statistics of these HL traits were characterized by inflation due to polygenicity (genomic-control lambda>1.09) and not because of possible confounders (LDSC intercept < 1.03; Supplemental Table 3). With respect to STR-derived traits, the SNP-h^2^ was lower <1% with only right-ear assessment with an estimate statistically different from zero (SRT-right SNP-h^2^= 0.009±0.002; Table 1). The limited informativeness of the SRT test in genetic studies is in line with previous findings^16, 53^. Accordingly, the SRT-derived traits were excluded for further evaluations.

**Table 1:**
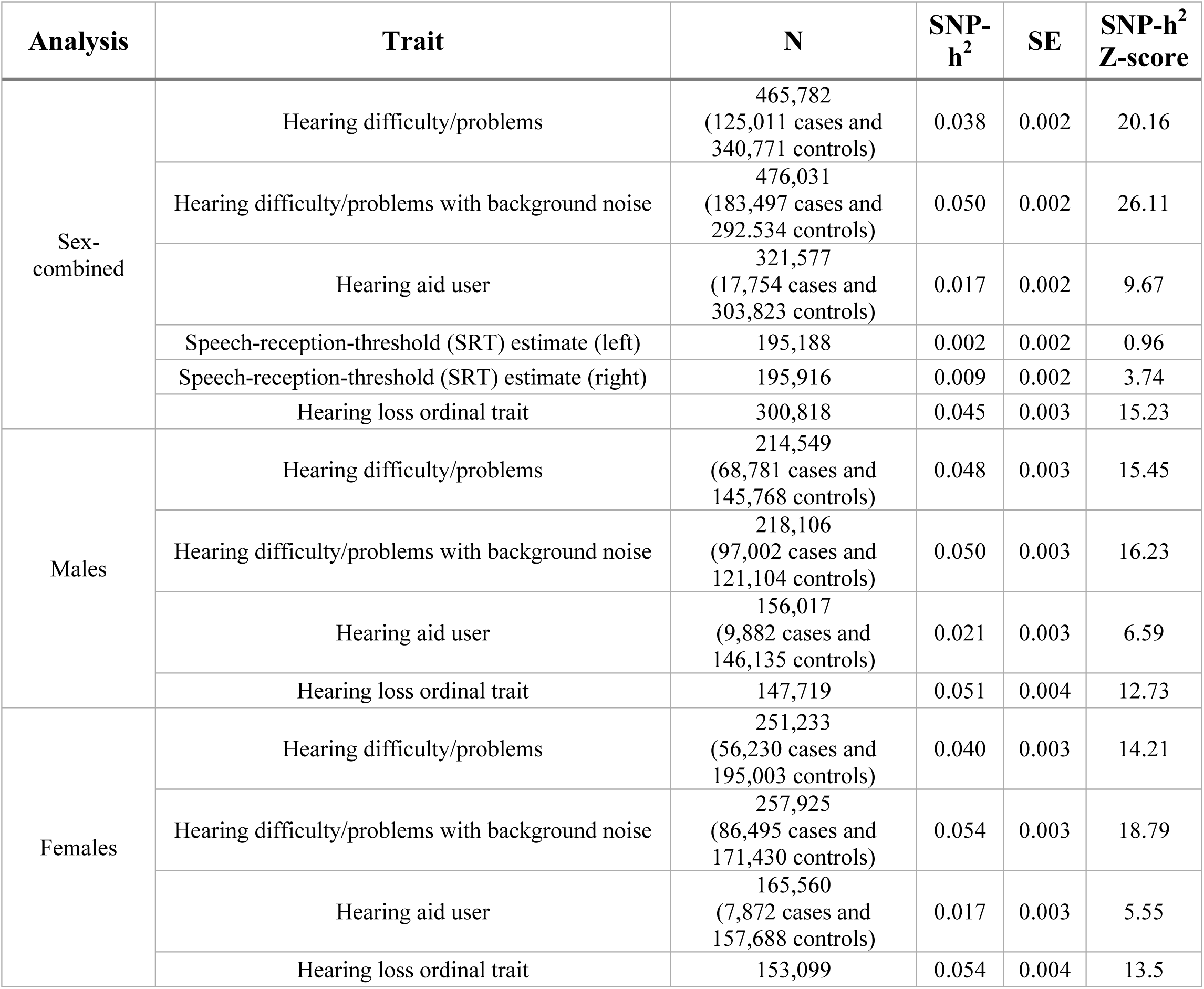
SNP heritability (SNP-h^2^) of hearing loss related traits assessed in the UK Biobank. SE: standard error.

| Analysis | Trait | N | SNP-h<sup>2</sup> | SE | SNP-h<sup>2</sup><br>Z-score |
| --- | --- | --- | --- | --- | --- |
| Sex-combined | Hearing difficulty/problems | 465,782<br>(125,011 cases and 340,771 controls) | 0.038 | 0.002 | 20.16 |
|  | Hearing difficulty/problems with background noise | 476,031<br>(183,497 cases and 292,534 controls) | 0.050 | 0.002 | 26.11 |
|  | Hearing aid user | 321,577<br>(17,754 cases and 303,823 controls) | 0.017 | 0.002 | 9.67 |
|  | Speech-reception-threshold (SRT) estimate (left) | 195,188 | 0.002 | 0.002 | 0.96 |
|  | Speech-reception-threshold (SRT) estimate (right) | 195,916 | 0.009 | 0.002 | 3.74 |
|  | Hearing loss ordinal trait | 300,818 | 0.045 | 0.003 | 15.23 |
| Males | Hearing difficulty/problems | 214,549<br>(68,781 cases and 145,768 controls) | 0.048 | 0.003 | 15.45 |
|  | Hearing difficulty/problems with background noise | 218,106<br>(97,002 cases and 121,104 controls) | 0.050 | 0.003 | 16.23 |
|  | Hearing aid user | 156,017<br>(9,882 cases and 146,135 controls) | 0.021 | 0.003 | 6.59 |
|  | Hearing loss ordinal trait | 147,719 | 0.051 | 0.004 | 12.73 |
| Females | Hearing difficulty/problems | 251,233<br>(56,230 cases and 195,003 controls) | 0.040 | 0.003 | 14.21 |
|  | Hearing difficulty/problems with background noise | 257,925<br>(86,495 cases and 171,430 controls) | 0.054 | 0.003 | 18.79 |
|  | Hearing aid user | 165,560<br>(7,872 cases and 157,688 controls) | 0.017 | 0.003 | 5.55 |
|  | Hearing loss ordinal trait | 153,099 | 0.054 | 0.004 | 13.5 |

The genetic correlations among the questionnaire-derived HL traits in UKB ranged from rg=0.830±0.013 between “*Hearing difficulties”* and “*Hearing difficulties with background noise*”, to rg=0.393±0.039 between “*Hearing difficulties with background noise*” and “*Hearing aid use*” (Figure 1; Supplemental Table 4).

**Figure 1:**
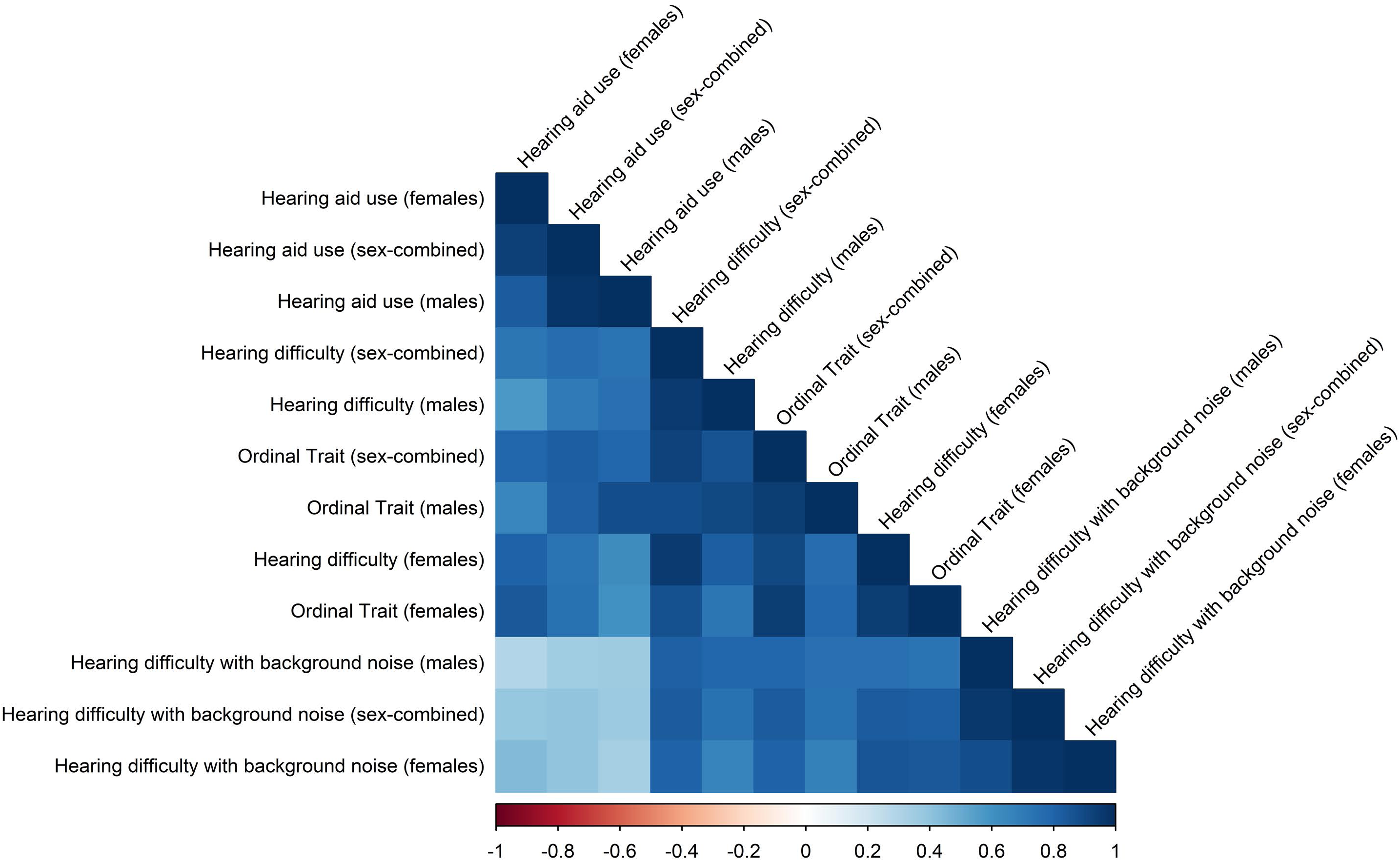
Genetic correlation among hearing loss traits assessed via questionnaire in UK Biobank. The square shade intensity is proportional to the magnitude of the correlation.

The ordinal phenotype derived from combining the three questionnaire-derived HL traits in UKB (Supplemental Table 1; hereafter abbreviated as HL-ORD) showed a high genetic correlation with each of the binary phenotypes, ranging from rg= 0.927±0.015 with “*Hearing difficulties”* to rg=0.830±0.020 with “*Hearing aid use*”.

To further investigate the differences across the questionnaire-derived HL traits, we conducted a sex-stratified analysis and tested the genetic correlation among whole-sample and sex-stratified analyses in UKB and also with respect to HL assessed in NHS (female participants only; SNP- h^2^=0.077±0.020) and HPFS (male participants only; SNP-h^2^=0.161±0.053). Considering each of the HL traits tested, no difference was observed between sexes in the SNP-h^2^ estimates (Table 1). Among the UKB questionnaire-derived HL traits, “*Hearing difficulties with background noise*” showed the highest rg between males and females (rg=0.882±0.035), while the lowest rg between sexes was observed for the HL-ORD trait (rg=0.782±0.050).

Considering the HPFS HL outcome and male-specific UKB analyses, the highest genetic correlation was observed for *“Hearing difficulty”* (rg=0.633±0.136). The same trait was the one with the highest genetic correlation with the NHS HL outcome among the UKB female-specific analyses (rg=0.737±0.115). Based on the genetic correlations among UKB, HPFS, and NHS, the UKB *“Hearing difficulties”* was defined as the primary UKB phenotype and the loci identified with respect to this trait were tested for single-variant and PRS replication using the NHS and HPFS datasets. Additionally, to maximize the discovery of our analyses, we also meta-analyzed UKB, NHS, and HPFS cohorts (278,744 female participants from UKB and NHS and 223,081 male participants from UKB and HPFS).

### Genome-wide association analyses

In UKB sex-combined analysis, we identified 46 LD-independent genome-wide significant (GWS) loci (P < 5×10^-8^) associated with hearing difficulties (Supplemental Table 5). In the sex-stratified analyses, we identified 14 and 19 LD-independent GWS loci in males and females, respectively (Supplemental Tables 6 and 7). In a few instances, the top-index variant identified in the sex-stratified analyses was LD-independent from the ones mapped in the GWAS loci observed in the entire cohorts. Additionally, we identified six variants with statistical differences in the sex-specific effects of the HL-associated variants (Supplemental Tables 6 and 7; rs2876317 in *PDE7B;* rs9784468 in *SORBS2*, rs1808828 in *ABLIM3*, rs9677089 in *SPTBN1*, rs12515096 in *ARHGEF28*, and rs35624969 in *TBL2*).

Beyond single-variant associations, the PRS derived from UKB were significantly associated with hearing difficulties in NHS and HPFS considering sex-combined and sex-stratified analyses (sex-combined PRS on NHS+HPFS: R^2^=0.05%, p=6.59×10^-6^; female-specific PRS on NHS: R^2^=0.47%, p=1.77×10^-30^, male-specific PRS on HPFS: R^2^=0.5%, p=2.84×10^-11^).

The GWAS meta-analysis of UKB, NHS, and HPFS (N=501,825) provided a higher statistical power, identifying 54 LD-independent loci significantly associated with hearing difficulties (Table 2).

**Table 2:** LD-independent loci associated with hearing difficulties reaching genome-wide significance (p<5×10^-8^) in the meta-analysis of UK Biobank (UKB), Nurses’ Health Studies (NHS; I and II), and Health Professional Follow-up Study (HPFS). The results of the replication analysis in Million Veteran Program (MVP) are also reported. Abbreviations: chromosome (CHR), position (POS); base pairs (bp); Effect Allele Frequency (EAF); Odds Ratio (OR), P value (P).

| rsID | CHR | POS (bp) | Effect Allele | Other Allele | UKB-NHS-HPFS meta-analysis |  | MVP Replication |  |
| --- | --- | --- | --- | --- | --- | --- | --- | --- |
|  |  |  |  |  | OR | P | OR | P |
| rs36062310 | 22 | 50,988,105 | A | G | 1.15 | $2.75 \times 10^{-32}$ | 1.08 | $4.80 \times 10^{-6}$ |
| rs6871548 | 5 | 73,076,024 | T | C | 1.05 | $2.30 \times 10^{-25}$ | 1.04 | $1.23 \times 10^{-8}$ |
| rs34993346 | 11 | 89,046,097 | A | G | 0.95 | $1.38 \times 10^{-24}$ | 1.00 | 0.872 |
| rs2254303 | 6 | 43,276,390 | A | G | 1.05 | $1.19 \times 10^{-23}$ | 1.04 | $5.23 \times 10^{-11}$ |
| rs10901863 | 10 | 126,812,270 | T | C | 1.05 | $6.21 \times 10^{-23}$ | 1.06 | $1.17 \times 10^{-11}$ |
| rs739138 | 22 | 38,122,462 | A | G | 0.95 | $4.70 \times 10^{-19}$ | 0.96 | $6.21 \times 10^{-8}$ |
| rs67307131 | 11 | 118,480,223 | T | C | 0.96 | $3.12 \times 10^{-18}$ | 0.97 | $2.83 \times 10^{-7}$ |
| rs9493627 | 6 | 133,789,728 | A | G | 1.04 | $1.03 \times 10^{-17}$ | 1.06 | $1.91 \times 10^{-19}$ |
| rs11073804 | 15 | 89,252,012 | A | T | 0.95 | $5.60 \times 10^{-17}$ | 0.91 | $3.50 \times 10^{-28}$ |
| rs6662164 | 1 | 46,146,230 | A | T | 1.04 | $6.59 \times 10^{-17}$ | 1.04 | $7.44 \times 10^{-9}$ |
| rs72622588 | 3 | 182,003,490 | T | G | 0.95 | $1.53 \times 10^{-16}$ | 0.97 | 0.002 |
| rs1566128 | 14 | 52,514,981 | A | G | 1.04 | $1.02 \times 10^{-15}$ | 1.04 | $3.98 \times 10^{-11}$ |
| rs6545432 | 2 | 54,817,683 | A | G | 1.04 | $1.05 \times 10^{-15}$ | 1.04 | $2.37 \times 10^{-8}$ |
| rs11041717 | 11 | 8,054,933 | A | C | 0.96 | $7.31 \times 10^{-15}$ | 0.98 | 0.071 |
| rs2877561 | 3 | 121,712,051 | A | C | 1.04 | $1.62 \times 10^{-14}$ | 1.03 | $4.95 \times 10^{-4}$ |
| rs13147559 | 4 | 17,524,570 | C | G | 0.95 | $3.81 \times 10^{-14}$ | 0.99 | 0.596 |
| rs2296506 | 6 | 158,507,981 | A | G | 1.03 | $1.70 \times 10^{-13}$ | 1.02 | $3.36 \times 10^{-4}$ |
| rs13171669 | 5 | 148,601,243 | A | G | 0.97 | $1.15 \times 10^{-12}$ | 0.98 | 0.007 |
| rs12156228 | 8 | 141,701,299 | T | G | 0.97 | $3.67 \times 10^{-12}$ | 0.97 | $6.31 \times 10^{-4}$ |
| rs222836 | 17 | 7,133,162 | A | G | 0.97 | $1.13 \times 10^{-11}$ | 0.98 | $7.37 \times 10^{-4}$ |
| rs4948502 | 10 | 63,839,417 | T | C | 1.03 | $1.26 \times 10^{-11}$ | 1.01 | 0.229 |
| rs11238325 | 7 | 50,853,151 | T | C | 1.04 | $1.88 \times 10^{-11}$ | 1.04 | $5.65 \times 10^{-6}$ |
| rs35094336 | 8 | 82,670,771 | A | G | 1.06 | $2.07 \times 10^{-11}$ | 1.03 | 0.021 |
| rs7712395 | 5 | 2,559,538 | T | C | 1.05 | $3.64 \times 10^{-11}$ | 1.04 | $6.45 \times 10^{-5}$ |
| rs143282422 | 10 | 73,377,112 | A | G | 1.16 | $4.43 \times 10^{-11}$ | 1.22 | $8.30 \times 10^{-11}$ |
| rs6675438 | 1 | 165,112,224 | A | T | 1.03 | $8.82 \times 10^{-11}$ | 1.03 | $1.62 \times 10^{-4}$ |
| rs57167368 | 10 | 80,521,351 | A | G | 0.96 | $9.91 \times 10^{-11}$ | 0.97 | 0.001 |
| rs72930998 | 18 | 52,632,968 | T | C | 0.97 | $2.03 \times 10^{-10}$ | 0.98 | 0.028 |
| rs1171114 | 6 | 84,227,646 | T | C | 0.97 | $2.69 \times 10^{-10}$ | 0.98 | 0.018 |
| rs2354376 | 7 | 138,487,145 | A | C | 1.03 | $3.17 \times 10^{-10}$ | 1.01 | 0.100 |
| rs717973 | 2 | 208,091,112 | A | G | 0.97 | $4.61 \times 10^{-10}$ | 0.99 | 0.072 |
| rs7819550 | 8 | 74,206,582 | A | G | 1.04 | $6.96 \times 10^{-10}$ | 1.05 | $3.92 \times 10^{-9}$ |
| rs72818515 | 10 | 76,060,718 | A | C | 1.03 | $7.08 \times 10^{-10}$ | 1.01 | 0.186 |
| rs112725535 | 1 | 6,494,086 | A | G | 0.96 | $7.85 \times 10^{-10}$ | 0.99 | 0.318 |
| rs11022697 | 11 | 13,178,285 | C | G | 0.97 | $1.17 \times 10^{-9}$ | 0.97 | $1.15 \times 10^{-5}$ |
| rs34073570 | 5 | 103,998,895 | CT | C | 0.97 | $1.32 \times 10^{-9}$ | 0.99 | 0.316 |
| rs61863078 | 10 | 94,777,108 | A | G | 1.03 | $1.54 \times 10^{-9}$ | 1.00 | 0.777 |
| rs111935448 | 16 | 30,919,807 | T | C | 1.04 | $1.75 \times 10^{-9}$ | 0.99 | 0.218 |
| rs58919600 | 5 | 92,970,519 | T | C | 1.04 | $1.86 \times 10^{-9}$ | 1.03 | 0.002 |
| rs11085064 | 19 | 4,209,152 | A | G | 1.04 | $2.84 \times 10^{-9}$ | 1.03 | $9.49 \times 10^{-4}$ |
| rs9783279 | 11 | 68,972,992 | A | C | 0.97 | $2.97 \times 10^{-9}$ | 1.00 | 0.872 |
| rs8102051 | 19 | 2,370,476 | A | C | 1.03 | $3.38 \times 10^{-9}$ | 1.01 | 0.429 |
| rs5800853 | 12 | 109,790,748 | CAG | C | 1.03 | $4.87 \times 10^{-9}$ | 1.02 | 0.017 |
| rs1928176 | 6 | 21,968,899 | A | G | 0.97 | $5.44 \times 10^{-9}$ | 0.99 | 0.028 |
| rs9530470 | 13 | 76,416,638 | A | G | 0.97 | $5.64 \times 10^{-9}$ | 0.99 | 0.051 |
| rs148512269 | 2 | 227,596,034 | T | G | 1.11 | $9.83 \times 10^{-9}$ | 1.05 | 0.133 |
| rs61734651 | 20 | 61,451,332 | T | C | 1.06 | $1.30 \times 10^{-8}$ | 1.03 | 0.028 |
| rs8063057 | 16 | 53,812,433 | T | C | 1.03 | $1.64 \times 10^{-8}$ | 1.01 | 0.081 |
| rs566673 | 11 | 66,401,373 | T | G | 0.97 | $1.80 \times 10^{-8}$ | 1.01 | 0.277 |
| rs78229182 | 6 | 151128784 | T | C | 1.05 | $2.07 \times 10^{-8}$ | 0.98 | 0.106 |
| rs73204028 | 7 | 115,092,852 | T | C | 0.97 | $2.20 \times 10^{-8}$ | 0.98 | 0.012 |
| rs766262445<br>(MVP LD-proxy: rs9536378) | 13 | 53,830,039 | CT(A) | C | 1.03 | $2.63 \times 10^{-8}$ | 1.00 | 0.787 |
| rs2273654 | 10 | 102,689,217 | T | C | 0.97 | $4.01 \times 10^{-8}$ | 0.99 | 0.425 |
| rs35624969 | 7 | 72,991,592 | T | C | 0.97 | $4.66 \times 10^{-8}$ | 0.99 | 0.218 |

In the sex-stratified investigation, we gained one additional locus in the male-specific GWAS meta-analysis (15 LD-independent GWS loci) and five additional loci in the female-specific GWAS meta-analysis (24 LD-independent GWS loci). Similar to the UKB-only GWAS, few index variants identified in the sex-specific GWAS meta-analysis were LD-independent of the loci identified in the overall GWAS meta-analysis and we identified statistical differences between sexes in their effect size for three variants (Supplemental Table 8; rs13399656 mapped to *SPTBN1*, rs1808828 in *ABLIM3*, and rs11738813 in *ARHGEF28)*.

Considering the 54 loci reaching genome-wide significance in the sex-combined meta-analysis, we tested whether the effects detected were replicated in an independent sample, 226,043 MVP participants of European descent. We observed that 34 loci were at least nominally replicated in the MVP cohort (Table 2; p<0.05). Considering concordance between discovery and replication cohorts, 49 loci showed consistent effect direction. The probability of observing concordant directions of 49 out of the 54 loci tested by chance is 5.9×10^-6^. Leveraging our discovery GWAS meta-analysis as a training dataset, we conducted a PRS analysis in the MVP cohort that showed highly significant prediction across all variant-inclusion thresholds tested (p<1.20×10^-103^; Supplemental Table 9).

To assess the cross-ancestry generalizability of the loci identified by the GWAS meta-analysis, we conducted a replication analysis of the single-variant associations in the UKB participants of non-European descent (AFR N=6,636; AMR N=980; CSA N=8,876; EAS N=2,709; MID N=1,599). Considering a nominal significance threshold, we replicated (p<0.05) the associations of four variants (rs6675438, rs61863078, rs2273654, and rs61734651) in AFR, five variants (rs666720, rs1220628, rs271143, rs9493627, rs146229052) in AMR, ten variants were replicated in CSA (rs666720, rs6662164, rs148512269, rs6869243, rs6871548, rs296430, rs2296506, rs766262445, rs72930998, and rs1005473), and two variants in EAS (rs11238325, and rs34993346) (Supplemental Table 10). The limited number of single-loci replication is due to the dramatic difference in sample size between the EUR discovery cohort (N=501,825) and the non-EUR replication cohorts (total N=20,800). However, we observed a significant cross-ancestry transferability in the HL PRS considering the GWAS meta-analysis and UKB-only GWAS as training datasets. Specifically, we observed an increased statistical power of the PRS derived from the GWAS meta-analysis (UKB-NHS-HPFS PRS: AFR R^2^=4.73%, p=4.65×10^-68^; AMR R^2^=1.96%, p=1.23×10^-5^; CSA R^2^=3.5%, p=6.26×10^-66^; EAS R^2^=2.45%, p=1.94×10^-15^) when compared to the one derived from the UKB-only GWAS (UKB-only PRS: AFR R^2^=0.14%, p=0.001; AMR R^2^=0.54%, p=0.0140; CSA R^2^=0.13%, p=6.01×10^-4^; EAS R^2^=0.08, p=0.076). Due to the limited sample size available, we did not conduct a sex-stratified cross-ancestry replication analysis with respect to single-variant and PRS associations.

Because of the higher statistical power, the following sections describe the *in-silico* analyses conducted on the basis of the GWAS meta-analyses of UKB (field ID 2247 “*Hearing difficulties*”), NHS (“*Hearing problems*”), and HPFS (“H*earing problems*”).

### Variant prioritization, fine-mapping, and multi-tissue transcriptome-wide association study

To translate genetic associations into information regarding potential causal genes associated with hearing difficulties, we integrated different approaches ranging from positional mapping to imputation of genetically-regulated transcriptomic variation.

First, we performed a fine-mapping analysis for each GWS locus. Based on the PIP of the 4,382 SNP associations reaching GWS in the sex-combined GWAS meta-analysis, we identified 218 variants that are most likely to be causal (PIP> 30%; Supplemental Figure 1). Considering a CADD score threshold of 10 (top 1% of pathogenic variants across the human genome), we further prioritized 24 variants that mapped to 18 unique genes (Supplemental Table 11). Some of them (e.g., *ARID5B, CTBP2,* and *FTO*) were previously identified as causal loci of Mendelian forms of deafness and HL^8, 16^. Applying the same mapping strategy to the sex-stratified GWS loci, we identified 84 and 46 variants in the credible set for female- and male-specific analyses, respectively (Supplemental Figures 2 and 3). These included six female-specific and two male-specific pathogenic variants (CADD score>10). The sex-stratified mapping genes mostly overlapped with the ones identified in the sex-combined GWAS meta-analysis.

In addition to the positional mapping approach, we also implemented an independent method based on the genetic regulation of transcriptomic variation. Combining tissue-specific information regarding expression quantitative trait loci (eQTL), we conducted a multi-tissue TWAS using the S-MultiXcan approach^47^. In the sex-combined meta-analysis, we identified 107 transcriptome-wide significant (TWS) genes (multi-tissue p<2.24×10^-6^; Figure 2; Supplemental Table 12).

**Figure 2:**
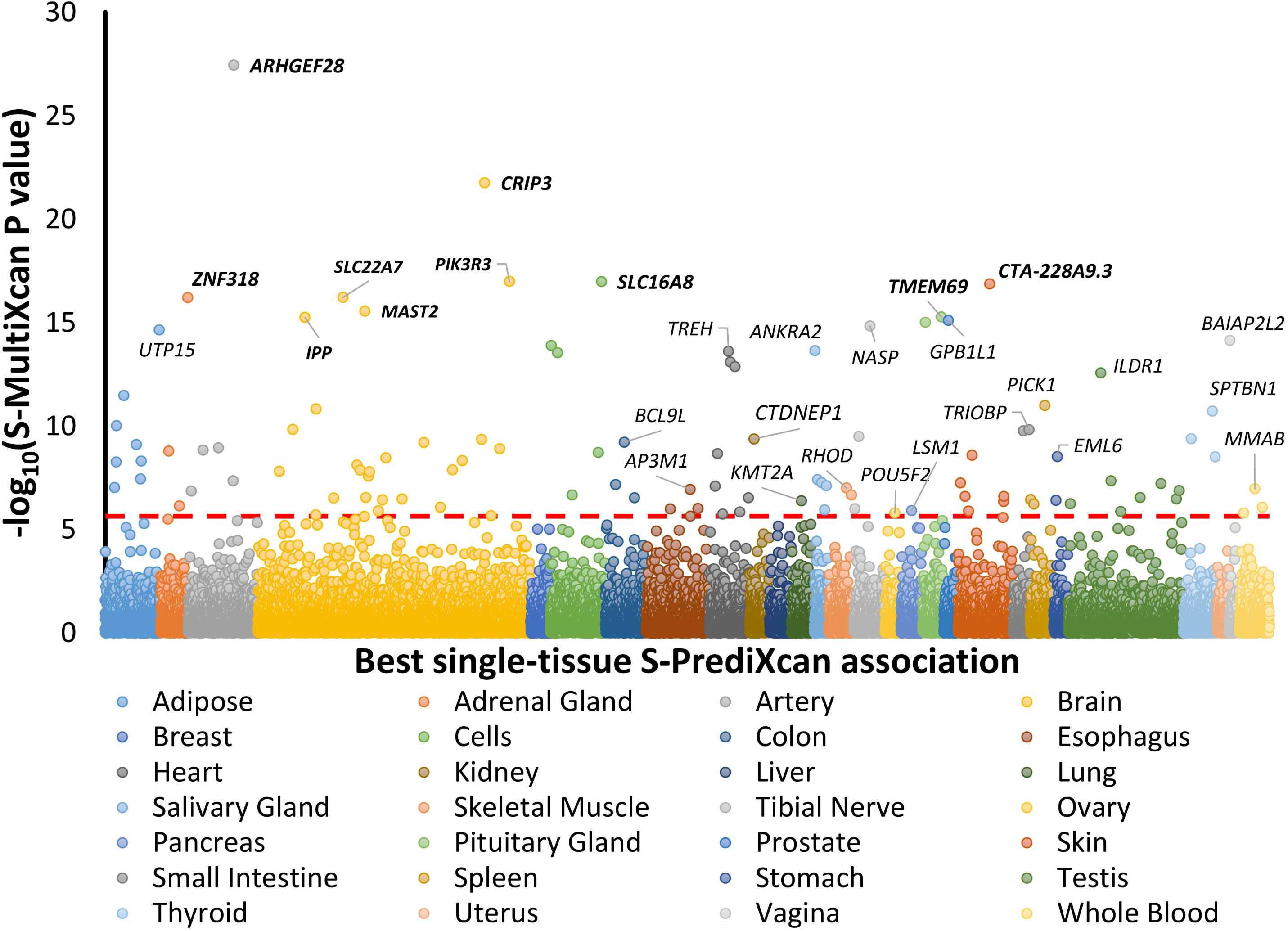
Multi-tissue transcriptome-wide association study of hearing loss based on the sex-combined meta-analysis. The y-axis corresponds to two-tailed −log10 (p-value of the S-MultiXcan association). The x-axis reports the genes grouped based on the best single-tissue S-PrediXcan association. The red line refers to the Bonferroni multiple testing correction accounting for the number of genes tested (p<2.24×10^-6^). Bold labels are reported for the top-10 Bonferroni significant association. Additional labels are included for the top significant result for each tissue. Detailed results are available in Supplemental Table 12.

Considering top tissue-specific effect underlying the cross-tissue associations, we observed that five of the top-10 strongest associations were brain-related: putamen basal ganglia (*CRIP3* p=1.19×10^- 23^), spinal cord c-1 (*PIK3R3* p=1.32×10^-19^), pituitary gland (*TMEM69* p=2.14×10^-18^), caudate basal ganglia (*IPP* p=3.04×10^-18^), cerebellar hemisphere (*SLC22A7* p=3.56×10^-18^), and cerebellum (*MAST2* p=3.17×10^-17^). The strongest non-brain related TWS associations included tibial artery (*ARHGEF28* p=3.29×10-17), cultured fibroblasts (*ACAN* p=5.17×10^-17^), left ventricle (*DLK2* p=1.03×10^-16^), and tibial nerve (*NASP* p=1.42×10^-16^). In the sex-stratified meta-analysis, we identified 55 and 26 multi-tissue TWS associations in females and males, respectively (Supplemental Table 13). The sex-specific loci mostly overlapped with those identified in the sex-combined TWAS: i) 14 genes were TWS in three analyses; ii) 45 out of 55 female-specific TWS genes were also significant in the sex-combined TWAS; iii) 23 out of the 26 male-specific TWS genes were also significant in the sex-combined TWAS. However, for tissue-specific effects, we observed that the top genes were not related to brain transcriptomic regulation (top female-specific association: liver-*CRIP3* p=1.59×10^-18^; top male-specific association: spleen-*PHLDB1* p=1.59×10^- 11^). Additionally, among female-specific associations, we identified associations related to female organs: mammary gland (*ABCC10* p=6.16×10^-15^) and vagina (*BAIAP2L2* p=7.45×10^-13^).

### Enrichment for regulatory elements and biological pathways

To dissect further its polygenic architecture, we partitioned HL SNP-h^2^ with respect to regulatory elements to uncover enrichment of relevant biological annotations and processes. We observed that several annotations related to the regulatory function of the human genome are more likely to be involved in HL predisposition than that expected by chance (Supplemental Table 14). These included evolutionary-conserved regions (e.g., “*genomic evolutionary rate profiling scores”* p=1.11×10^-16^) and –elements involved in epigenetic and transcriptomic regulation (e.g., “*CpG dinucleotide content*” p=1.53×10^-10^; “*super-enhancer regions*” p=1.54×10^-7^). In a complementary approach, we performed gene set enrichment analysis of HL-associated genes (based on positionally mapping). Two biological pathways were observed to be statistically overrepresented: “*response to trabectedin*” (p=1.58×10^-8^) and “*sensory perception of mechanical stimulus*” (p=1.69×10^-7^). With respect to both analyses, we did not identify statistical differences in the enrichments calculated from the sex-specific genetic associations.

### Phenome-wide genetic correlation and latent causal variable analysis of hearing loss

To investigate the genetic overlap of HL with other traits and diseases, we assessed phenome-wide genetic correlation considering 5,337 phenotypes for the sex-combined analysis, 2,353 for the female-specific analysis, and 2,249 for the male-specific analysis. Considering a Bonferroni correction accounting for the number of phenotypes tested in sex-combined, female, and male analyses, we identified 309, 79, and 109 significant genetic correlations, respectively (Figure 3; Supplemental Table 15).

**Figure 3:**
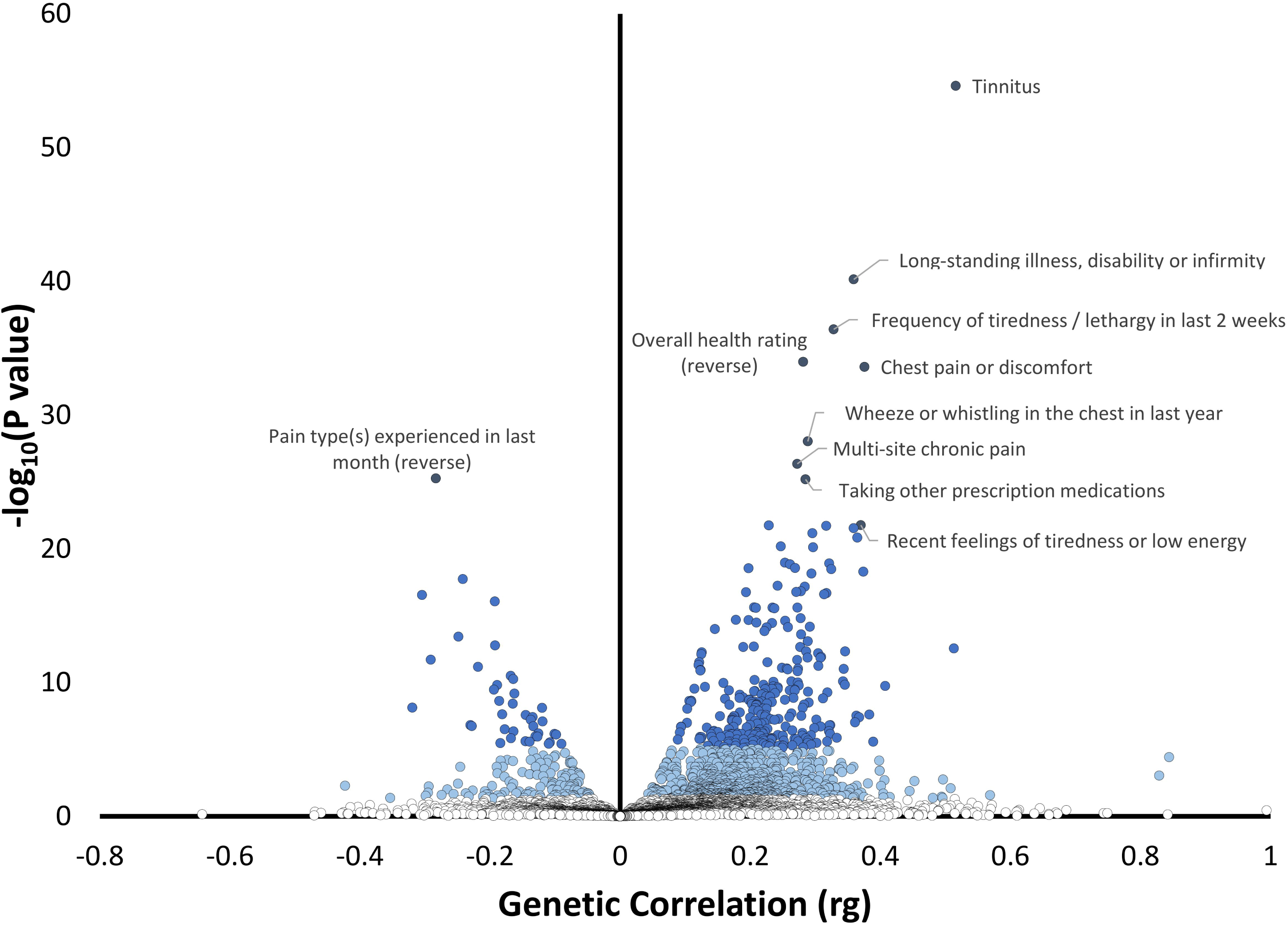
Phenome-wide genetic correlation of hearing loss in the sex-combined analysis. The x-axis reports the genetic correlation of hearing loss with the traits tested. The y-axis corresponds to two-tailed −log10(p-value). Blue shades correspond to significance strength, from white, non-significant (p>0.05), to light blue (nominal significance p<0.05), to blue (Bonferroni correction p<9×10-6), and dark blue (top 10 results). Phenotype labels are included for the top 10 results. Full results are reported in Supplemental Table 15.

With respect to the sex-combined GWAS meta-analysis, the strongest genetic correlation was “*Tinnitus*” (rg=0.52, p=2.44×10^-55^). Other strong positive genetic correlations included *“Long-standing illness, disability or infirmity”* (rg=0.36, p=7.12×10^-41^); and *“Frequency of tiredness / lethargy in last 2 weeks”* (rg= 0.33, p=3.85×10^-37^). Among negative HL genetic correlations, we observed *“Leisure/social activities”* (rg=-0.19, P=3.30×10^-10^) and “*Belief that own life is meaningful”* (rg=-0.18, p=3.38×10^-6^). Although many of the sex-specific genetic correlations overlapped with those shared with sex-combined analysis, we also identified five traits with statistically significant sex differences in their genetic correlation with HL. Four of them were related to educational attainment with the strongest one observed for *“Qualification: College or University degree”* (female rg=0.11, P=2.52×10^-5^, male rg=-0.12, P=1.15×10^-5^; p_sex-difference_=1.16×10^-9^). The fifth genetic correlation was related to *“Time spend outdoors in summer”* (female rg=-0.16, P=2.73×10^-5^, male rg=0.06, P=0.08; p_sex-difference_=1.95×10^-5^).

To distinguish genetic correlations due to shared genetic mechanisms from those due to possible cause-effect relationships, we conducted an LCV analysis, identifying 22 significant putative causal effects (p<5.68×10^-5^; Figure 4, Supplemental Table 16).

**Figure 4:**
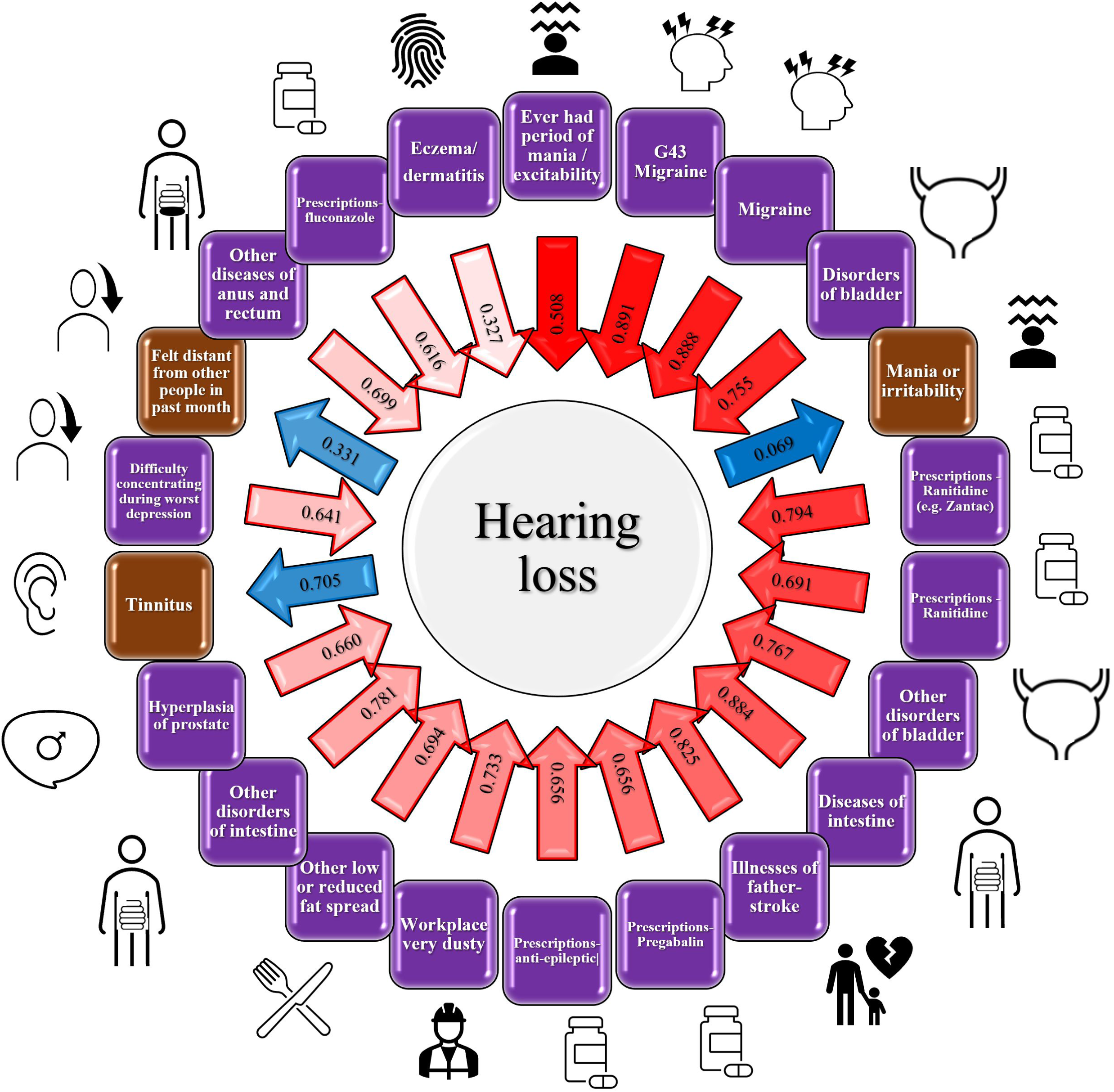
Visual representation of the 22 significant putative causal effects identified through the latent causal variable analysis. Brown labels: HL has causative effect for the trait in the label; Purple labels: Trait in the label has causative effect for HL. The absolute gcp (genetic causality proportion) value for each association is reported within the arrow, and the directions refer to the cause-effect relationship (Blue: HL causes Trait; Red: Trait causes HL). The shade intensity of the arrows is proportional to the statistical significance of the gcp estimates. Description of each trait tested, and details of the associations is available in Supplemental Table 16.

The traits with putative causal effect on HL included psychiatric traits (e.g., *“Ever had period of mania / excitability”* gcp=0.508, p=3.59×10^-23^), neurological disease (e.g., “*Migraine*” gcp=0.891, p=7.1×10^-21^), gastrointestinal outcomes (e.g., *“ICD-10 K63 Other diseases of intestine*” gcp=0.884, p=2.25×10^-11^), urogenital outcomes (e.g., *“ICD-10 N32 Other disorders of bladder*” gcp=0.755, p=7.83×10^-15^), medication use (e.g., *“Ranitidine*” gcp=0.691, p=1.38×10^-12^), cardiovascular diseases (“*Stroke family history*” gcp=0.825, p=6.67×10^-11^), immunological conditions (*“Eczema/dermatitis*” gcp=0.327, p=3.82×10^-5^), and workplace environment *(“Workplace very dusty”* gcp=0.733, p=2.04×10^-10^). We also identified genetic evidence that support a putative causal role on HL for four outcomes: *“Tinnitus*” (gcp=0.705, p=4.85×10^-8^); *“I was easily distracted”* (gcp=0.069, p=8.44×10^-15^), and *“Felt distant from other people in past month”* (gcp=0.331, p=6.52×10^-6^). Although it did not survive multiple testing correction, the strongest sex difference was observed with respect to *“Not having bipolar or depression disorder*” where there was a negative genetic causal effect on HL in females but not in males (females gcp= -0.73, p=0.005; males gcp=-0.28, p=0.147; p_sex-difference_=7.69×10^-4^).

### Multivariate gene-by-environment genome-wide interaction analysis

To investigate the role of gene-environment interactions in the predisposition to HL, we conducted a multivariate GEWIS. Because of the assumptions of the linear mixed model approach used^50^, we conducted this analysis with respect to the HL-ORD trait (UKB N=300,818). We tested 14 environmental factors simultaneously (Supplemental Table 2). In addition to sex, these include environments related to noise pollution (N=5) and tobacco smoking habits (N=8). Although no variant survived genome-wide testing correction, we identified 1,278 LD-independent nominally significant multivariate interactions (Supplemental Table 17). To identify environment-specific effects underlying the multivariate interactions of each of these variants, we considered BF>1 following StructLMM reccomendations^50^. We then conducted a GO enrichment analysis stratifying the LD-independent loci with nominally significant multivariate interactions based on the environment where they showed a BF>1. Considering FDR multiple testing correction at 5%, we identified 204 significant enrichments with respect to 57 unique GO terms. Most GO enrichments were shared across loci showing interaction effects with specific sex, noise, and smoking environmental factors (BF>1). Specifically, loci with environment-specific interactions related to sex, noise, and smoking showed 17 shared GO enrichments with respect to (e.g., GO:0007399 Nervous system development: “S*ex”* p=2.0×10^-6^; *“Average evening sound level of noise pollution”* p=1.3×10^-6^; *“Maternal smoking around birth”* p=3.6×10^-7^). Similarly, loci interacting only with noise and smoking environment (but not sex) shared 17 GO enrichments (e.g., GO:0030182 Neuron differentiation: *“Noisy workplace”* p=4.4×10^-5^; *“Ever smoked”* p=4.4×10^-6^) and two GO enrichments related to loci with environment-specific interactions related to sex and smoking (e.g., GO:0050767 Regulation of neurogenesis: “*Sex”* p=7.5×10^-5^; *“Maternal smoking around birth*” p=4.6×10^-5^). We also identified enrichments related to loci interacting only with a specific environment category: seven GO terms related to sex interactions (e.g., GO:0030857 Negative regulation of cell differentiation: *“Sex”* p=8.2×10^-5^), 10 GO terms related to noise-pollution interactions (e.g., GO:0050885 Neuromuscular process controlling balance: *“Noisy workplace”* p=3.7×10^-4^), and three GO terms related to smoking interactions (e.g., GO:0016101 Diterpenoid metabolic process: *“Smoking/smokers in household”* p=2.1×10^-4^).

## DISCUSSION

We conducted a large-scale investigation integrating information from genome-wide associations with tissue-specific transcriptomic variation and a causal inference analysis to translate genetic findings into insights regarding HL biology and epidemiology. Additionally, we expanded the focus of our investigation to differences between sexes and ancestries and to the interplay of genetic variation with noise pollution and tobacco smoking, two well-known HL risk factors.

Initially, we compared the SNP-h^2^ of different HL definitions available from UKB. These included three questionnaire-derived self-reported hearing traits and one trait derived from audiometric measures. Consistent with previous studies^16, 53^, we observed that SRT-derived assessment had a very low SNP-h^2^ (<1%) that did not permit us to investigate further this HL outcome. Conversely, self-reported HL outcomes showed significant SNP-h^2^ that was informative to investigate their polygenic architecture. We observed a high genetic correlation (rg>0.7) between them with the only exception being “*Hearing difficulty with background noise*” and “*Hearing aid use*” where the genetic correlation was much lower (rg<0.4). This may be because the proportion of individuals who seek hearing aids for their hearing loss is very low, even in the UK where hearing aids are fully covered by the National Health Service. Considering the HL outcome available in NHS and HPFS cohorts, we observed the highest correlation with the UKB outcome “*Hearing difficulties*”. Accordingly, these traits were used for the GWAS meta-analysis and the subsequent *in silico* investigations. These self-reported traits are considered relatively reliable instruments to investigate “real world” hearing impairment, although potentially influenced by psycho-social factors^16^. In the sex-combined GWAS meta-analysis including 501,825 individuals, we identified 54 LD-independent genome-wide significant associations, 14 of which are novel with respect to those identified by previous HL GWAS^8, 13, 16, 54^. In the UKB-NHS-HPFS meta-analysis, three variants showed statistical differences between sexes in their effect size. Specifically, rs13399656 in *SPTBN1* was the top findings in the sex-difference associations. This gene was previously recognized as a target for β-estradiol as the top upstream regulator^55^ and was already associated with altered expression in estrogen-responsive tissues in HL^8^. Similarly, rs1808828 mapped to *ABLIM3* and rs11738813 in *ARHGEF28* were identified as intronic variants mapped in genes involved in estrogen signaling pathway^56, 57^. These sex-specific associations changes in significant loci involved in hormonal regulating pathways may reflect the potential role of estrogen on hearing functions.

Although this primary analysis was conducted in individuals of European descent, we replicated several associations in an independent sample of 20,800 UKB participants of AFR, AMR, CSA, EAS, and MID descent. Additionally, we demonstrated that HL polygenic risk is partially shared across ancestry groups. However, consistent with previous studies of other health outcomes^58–60^, there are large differences in the predictive power due to the genetic diversity across human populations. Additionally, in some cases the HL variance explained by the cross-ancestry PRS association is larger than the one observed in the same-ancestry PRS association. This can be explained by two factors: i) the cross-ancestry results are based on the UKB cohort where the participants share the same HL assessment and sample characteristics; ii) the different LD structures of the ancestries investigated may have inflated the results observed.

The fine-mapping of the risk loci led to identifying putatively pathogenic variants (CADD score>10) mapped to genes previously demonstrated to be involved in HL pathogenesis. Specifically, we identified the exonic variant rs13147559 mapping in *CLRN2* as the strongest finding (CADD=22.5). This locus has been identified as causative for non-syndromic HL^61^ and may have a role in hair cell mechano-transduction^62^. Two exonic mutations in *NOL12:TRIOBP* were also identified as potentially deleterious (rs5756795 and rs7284476). *TRIOBP* alterations are related to both congenital deafness^63^ or Mendelian HL^15^. Mutations mapped to this gene have been thoroughly analyzed, especially in patients with a post-lingual HL (i.e., HL occurring after the development of normal speech)^64, 65^. Non-coding regulatory variants were also identified in HL-related genes such as *DLG4, ARID5B,* and *CTBP2.* In mice, elevated Arid5b in the aged cochlea is known to be critical for the etiologies of sensorineural HL, such as age-related HL, noise- and ototoxic drug-induced HL^66^. Conversely, *DLG4* was previously identified as significantly related to hearing due to its association with the hippocampal glutamatergic synapse pathways^67^. The deleterious variant mapping at *CTBP2* also supports the role of the central nervous system in HL pathogenesis. Indeed, CtBP2 is a marker for cochlear ribbon synapses, and deleterious variants (such as rs183893500, CADD=15.24) could influence vulnerability to cochlear synaptopathy (i.e., loss of nerve connections between the sensory cells and the central nervous system) in acquired sensorineural HL^68, 69^.

Our TWAS further highlighted that HL genetic basis is partially linked to brain transcriptomic regulation. We identified that several genes previously implicated in HL pathogenesis because of their potential involvement in the peripheral structures of the auditory system, which also showed predicted expression differences in various brain regions. For example, while *CRIP3* has broad expression in inner and outer hair cells^8^, we observed that transcriptomic changes of this gene in the putamen basal ganglia were associated with HL. Similarly, other genes that may be implicated in HL pathogenesis through both peripheral and central mechanisms include *IPP* (previously reported as highly expressed in human cochlear, cochlear hair cells, and spiral ganglion cells in mice^54^), *PIK3R3* (previously identified as specific to cell types in the cochlea^70^), *MAST2* (showing differential expression in the development of the inner ear^54^), and *SLC22A6* (identified as related to cochlear impairment^71^). Taken together, these results suggest that auditory dysfunction is not restricted to peripheral auditory structures, but subcortical neuroplastic changes involving the area receiving ascending signals from inferior brainstem nuclei could also be involved^72^. Additionally, several TWAS associations were related to the transcriptomic regulation in the cerebellum in line with the involvement of this brain region in auditory pathways^73^.

The sex-stratified TWAS showed several additional associations related to peripheral tissues. In particular, the female-specific TWAS identified as top findings transcriptomic changes of genes in breast mammary tissue (*ABCC10*) and vagina (*BAIAP2L2*). These associations in hormonally regulated tissues may reflect a potential role of estrogen in hearing function^74^. Indeed, our sex-stratified analyses identified associations mapped in genes involved in the estrogen signaling pathway (i.e., *SPTBN1*, *ABLIM3*, and *ARHGEF28*). With respect to the loci identified by the female-specific TWAS, *BAIAP2L2* is particularly interesting. In mice, mutations of the homologous gene Baiap2l2 were associated with alterations in hair cell transduction and deafness^75^ while the human locus *BAIAP2L2* is associated with suppression of the estrogen-mediated S–phase entry pathway in cell-cycle^76^. As the HL is associated with reduced estrogens levels, we hypothesize that the interplay between *BAIAP2L2* transcriptomic regulation and estrogen levels may play a role in HL in women.

We leveraged the genome-wide association statistics generated by our sex-combined and sex-stratified analyses to explore HL polygenic architecture in the context of other traits and diseases. Our phenome-wide genetic correlation analysis identified a wide range of health outcomes that share a significant proportion of their genetic liability with HL. The subsequent genetically-informed causal inference analysis showed that some of these genetic correlations may be due to cause-effect relationships linking HL to different health domains, including neurological, cardiovascular, and cancer-related outcomes. In line with the shared involvement of the auditory system, the strongest HL genetic correlation was with tinnitus (i.e., the conscious perception of an auditory sensation in the absence of a corresponding external stimulus^77^). This is consistent with the strong association between tinnitus and HL in the elderly^78^. A previous Mendelian randomization analysis showed a bidirectional relationship between them^28^. However, our LCV analysis supports that HL may have a causal effect on tinnitus. Further studies will be needed to understand the underlying dynamics of these comorbidities.

In addition, we observed several traits related to the psychological distress generally observed in elderly individuals. They included positive genetic correlations with disability/infirmity and tiredness/lethargy and negative genetic correlations with the propensity to social activities and the belief that life is meaningful. These findings illustrate that HL is associated with multiple aspects of quality of life in aging populations. We also identified five traits that showed statistically significant sex differences in their correlation with HL. Four of them were related to educational attainment where the genetic correlation with HL was positive in females and negative in males. A previous study demonstrated that the characteristics and the recruitment strategy of the UKB cohort (the largest sample in our meta-analysis) influenced some of the sex differences observable in this study population^79^. Accordingly, this sex difference may be specific to the structure of the UKB cohort. The other sex difference was related to time spent outside that showed a negative genetic correlation in females but not in males. This could be related to the higher comorbidity of HL with depression observed in women compared to men^80^.

Finally, our multivariate GEWIS analysis explored the interplay of genetic variation with sex differences and two established HL risk factors, noise pollution and tobacco smoking. Although no single interactive locus remained statistically significant after multiple testing correction, we observed several biological processes that are potentially involved in the inter-individual susceptibility to the effects of HL environmental risk factors. Loci interacting with sex, noise, and smoking environmental factors were mostly enriched for the same molecular pathways, suggesting that HL environmental risk factors may affect the same biological processes that lead to the onset of the diseases. In particular, we observed an overrepresentation of pathways related to brain developmental processes. An intriguing finding was the observed higher risk of HL with maternal smoking around birth, suggesting that early life exposures may influence later life auditory function.

Although our findings advance our understanding of HL and its consequences in adults, we also acknowledge several limitations. Similar to previous HL GWAS^8, 13–16^, our primary analyses were based on self-reported data. These appear to be more informative than the audiometric measures derived from SRT test, but they can still be biased by misreporting linked to cognitive processes, social desirability, and survey conditions. Our study was focused on acquired HL in adults and we excluded individuals with congenital HL when possible. However, we did not have information regarding HL age of onset across all cohorts investigated. Accordingly, a small proportion (<1%) of congenital HL cases may be present in our study populations. No sex-specific TWAS models are currently available from GTEx. Accordingly, our sex-stratified TWAS was conducted using sex-stratified genome-wide association statistics and sex-combined transcriptomic reference panels. This has likely reduced the statistical power of our analysis. Additionally, GTEx does not include tissues related to peripheral auditory system, limiting our ability to explore the differences in transcriptomic changes in the peripheral and central auditory system. Although we investigated cross-ancestry transferability of HL genetic risk, the limited diversity of the cohorts investigated did not permit us to explore the genetic basis of HL across ancestry groups. Similar to what observed for other complex traits, HL PRS explained a small phenotypic variance when applied across cohorts (e.g., the PRS derived from UKB-NHS-HPFS meta-analysis and applied to MVP sample).

In conclusion, we conducted a comprehensive investigation of the polygenic architecture of HL that i) identified novel risk loci, ii) provided evidence of the shared HL pathogenesis across human populations, iii) integrated genetic and transcriptomic data to dissect HL biology, iv) leveraged genome-wide information to explore the mechanisms underlying HL comorbidities, and v) uncovered possible biological processes that could underlie inter-individual differences in susceptibility to the effects of HL environmental risk factors. Further studies will be needed to translate this information into tools useful to improve HL preventive and therapeutic strategies.

## Data Availability

All data produced in the present work are contained in the manuscript.

## ACKNOWLEDGEMENTS

This research is funded by the National Institute on Deafness and Other Communication Disorders (R21 DC018098, UO1 DC010811). The authors also acknowledge grants from the National Institute on Drug Abuse (R33 DA047527), the National Institute on Mental Health (F32 MH122058), the National Cancer Institute (UM1 CA186107, UO1 CA167552, and UO1 CA176726), the One Mind, and the Marie Sklodowska-Curie Actions (Individual Fellowship 101028810). This research was performed using the UK Biobank Resource under the application 58146. This research is also based on data from the Million Veteran Program, Office of Research and Development, Veterans Health Administration, and was supported by award #MVP010. This publication does not represent the views of the Department of Veteran Affairs or the United States Government. We thank all the participants in the UK Biobank, the Nurse’s Health Studies, the Health Professionals Follow-up Study, and the Million Veteran Program.

## COMPETING INTERESTS

Dr Clifford reported receiving personal fees from Decibel Therapeutics outside the submitted work. Dr. S. Curhan serves as a consultant to Decibel Therapeutics. Dr. G. Curhan serves as a consultant to Decibel Therapeutics, AstraZeneca, Shire, Allena Pharmaceuticals, RenalGuard, OrfanBiotech, OM1, and Merck. He receives royalties from UpToDate for being an author and Section Editor. None of the other authors declare any competing interests.

## Figure Legends

**Supplemental Figure 1:** Visual representation of the fine-mapping analysis for sex-combined meta-analysis. Each panel refers to a GWS risk locus. The variants were identified according to their LD with respect to the lead SNP and inclusion in the credible set with at most ten causal variants.

**Supplemental Figure 2:** Visual representation of the fine-mapping analysis for female meta-analysis. Each panel refers to a GWS risk locus. The variants were identified according to their LD with respect to the lead SNP and inclusion in the credible set with at most ten causal variants.

**Supplemental Figure 3:** Visual representation of the fine-mapping analysis for male meta-analysis. Each panel refers to a GWS risk locus. The variants were identified according to their LD with respect to the lead SNP and inclusion in the credible set with at most ten causal variants.

## Notes

### Author Declarations

The use of UKB individual-level data has been conducted through the application reference no. 58146. UKB has approval from the North West Multi-center Research Ethics Committee (MREC) as a Research Tissue Bank (RTB) approval. This approval means that researchers do not require separate ethical clearance and can operate under the RTB approval. The use of MVP individual-level data was conducted under project #MVP010 approved by the VA Central IRB. The use of data from the Nurses' Health Studies and Health Professional Follow-Up Study has been approved by the Partners Ethics Board.

