## Supplementary figures and images for "Sex differences, cross-ancestry generalizability, and noise-smoking interactions in the polygenic architecture of hearing loss in adults"

### Supplemental Figure 2.png

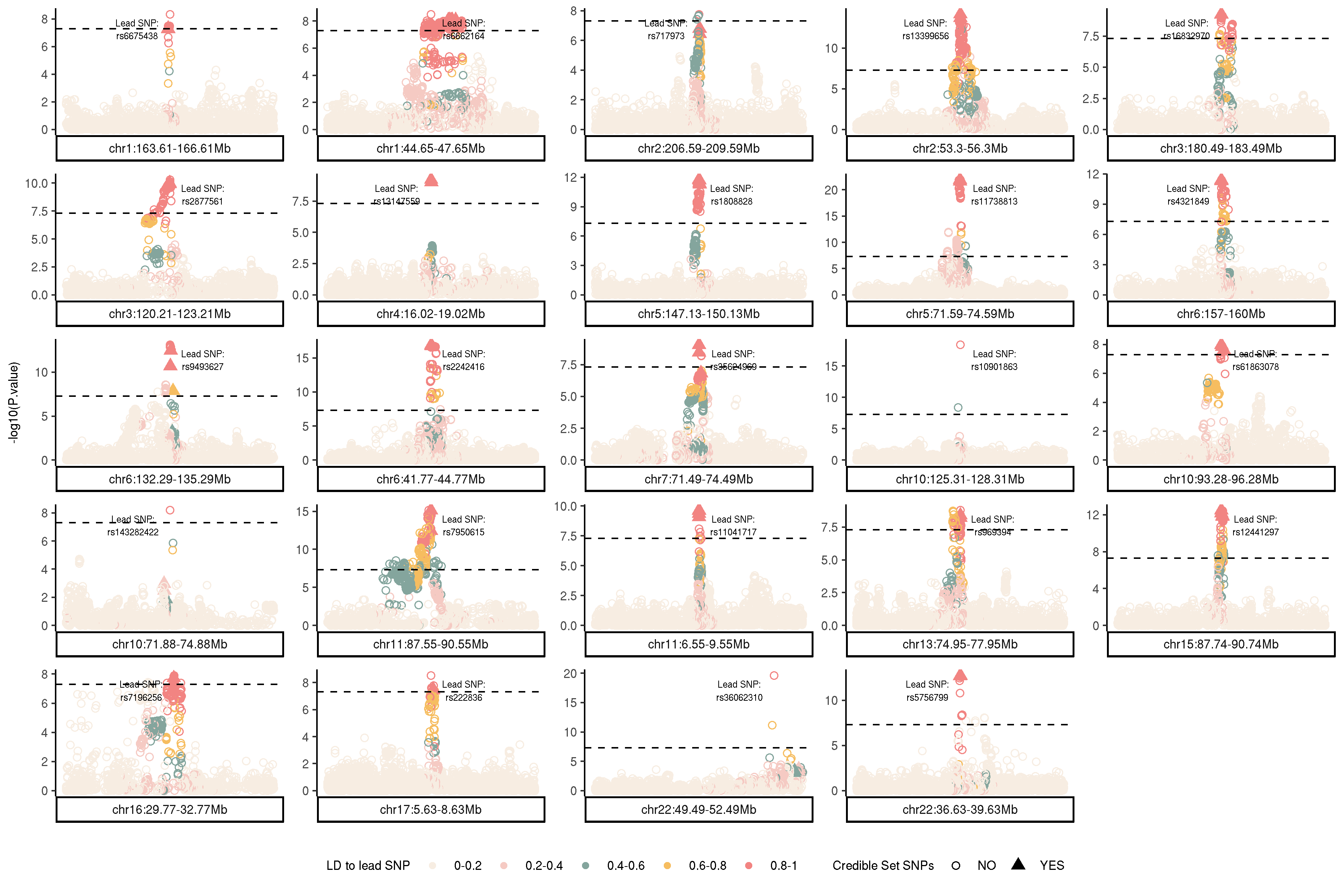

### Supplemental Figure 3.png

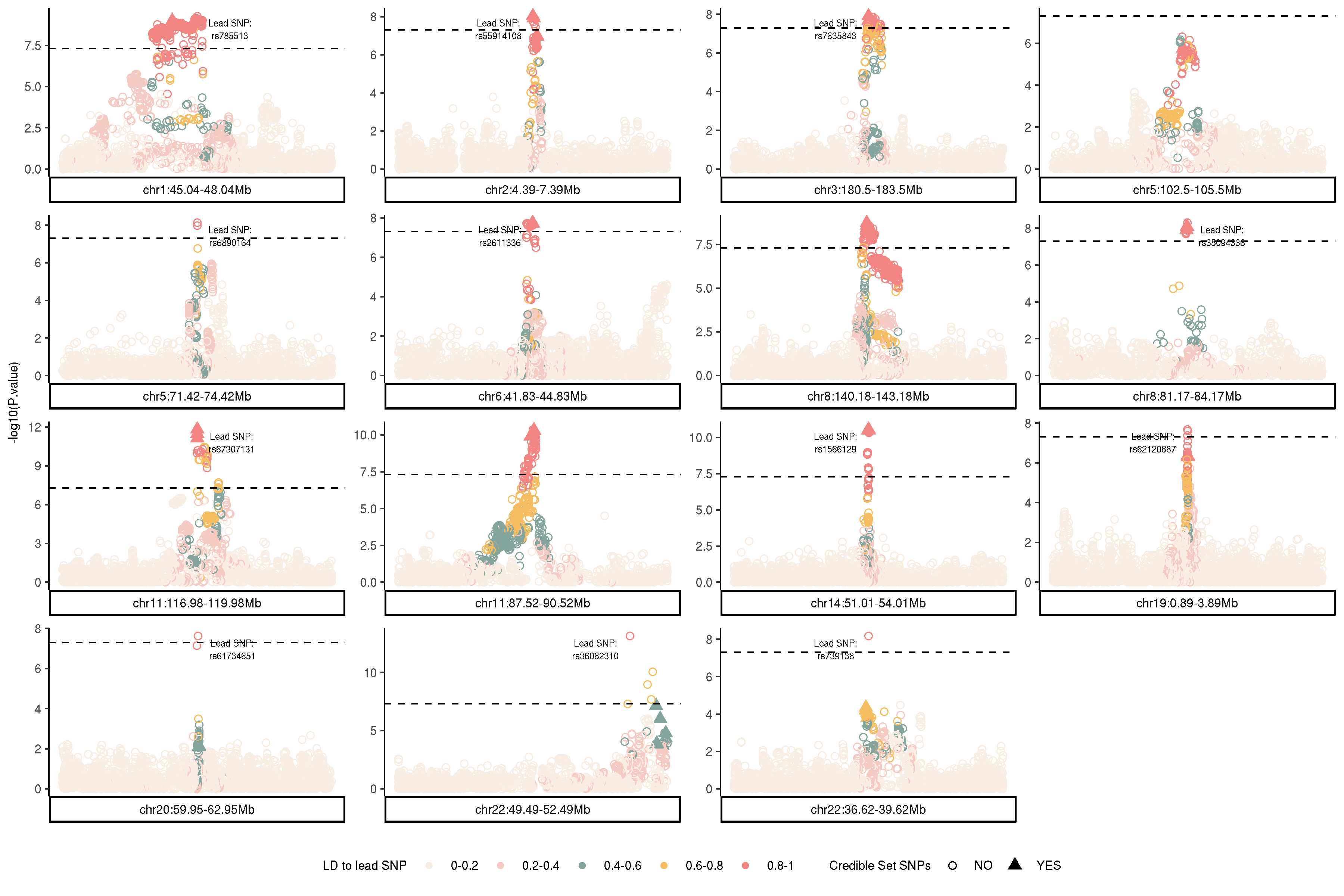

### Supplemetnal Figure 1.png

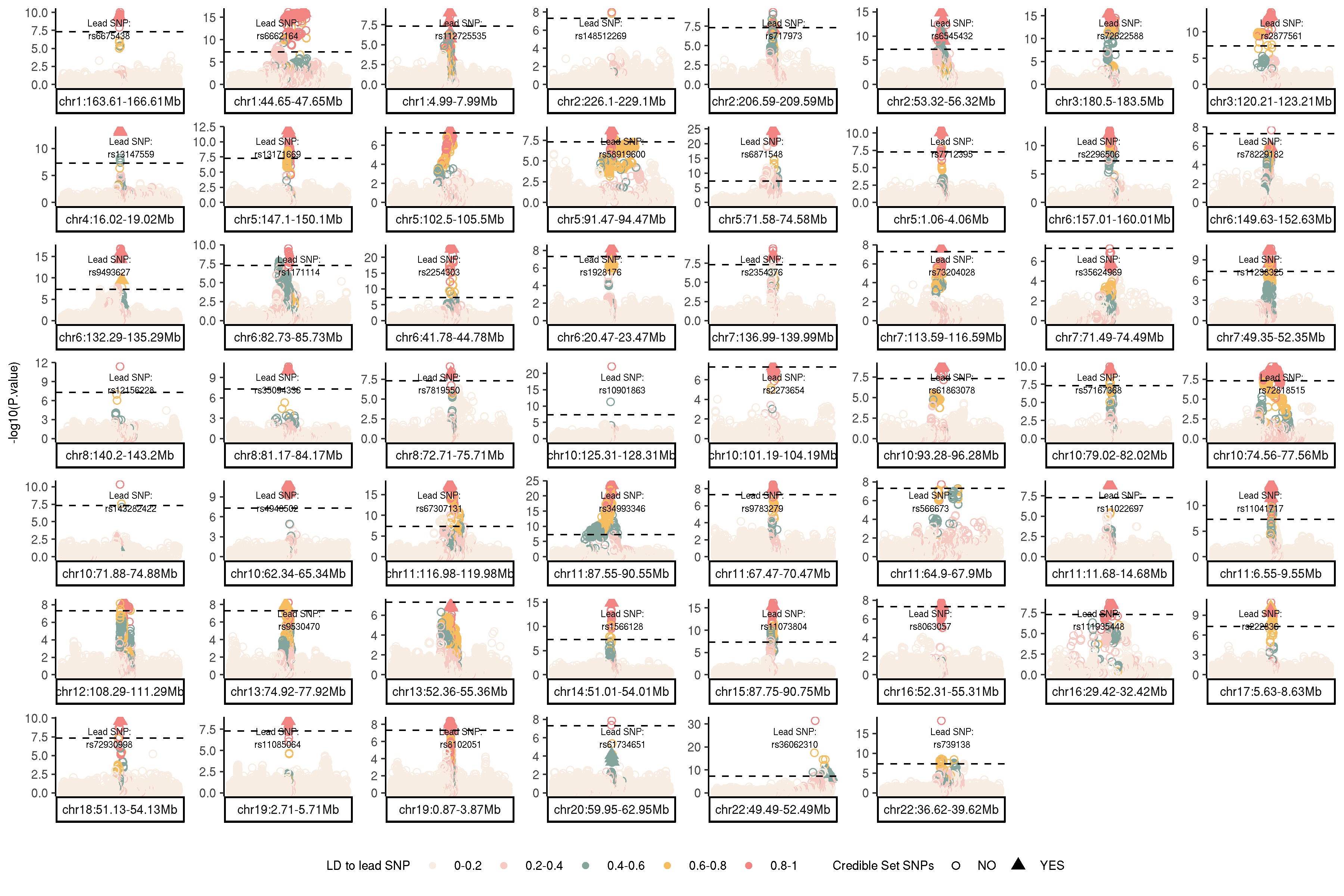
